# Structural-Fiscal Alignment in German Hospital Reform: Network Analysis of Service Group Prerequisites and Comparative Policy Scenarios

**DOI:** 10.64898/2025.12.01.25341348

**Authors:** Cornelius J. Werner, Michael Denkinger

**Author notes:** Corresponding author: Cornelius J. Werner, MD Johanniter-Hospital Stendal Wendstrasse 30, 39576 Hansestadt Stendal Germany.

## Abstract

**Background:** Germany’s Hospital Care Improvement Act (Krankenhausversorgungsverbesserungsgesetz, KHVVG) establishes 65 hospital service groups (Leistungsgruppen, LGs) with codified structural prerequisites, creating a directed dependency network. It remains unclear whether this regulatory structure appropriately reflects the economic importance of different services, particularly for specialties serving aging populations.

**Methods:** We modeled KHVVG dependencies as a directed network and calculated centrality metrics for all 65 LGs. Using elastic net regression, we related network position to LG-specific revenue data (2021) as a proxy for real-world service importance. Residuals between actual and network-predicted revenue quantified structural-fiscal alignment. We compared three policy scenarios: baseline KHVVG structure, KHAG reform (eliminating infectious diseases and emergency medicine service groups), and a hypothetical GBA scenario (adding geriatrics as mandatory prerequisite for hip replacement services).

**Results:** The regression model explained 56.6% of revenue variance (R²=0.566). Enablement index (β=+0.38) and weighted in-degree (β=−0.84) were primary predictors. Twenty-one LGs (35.6%) showed good alignment (|residual| <0.5 log units), including high-revenue services such as general internal medicine (€9.8B, residual +0.03) and general surgery (€12.1B, residual −0.06). Geriatrics (LG56) exhibited the largest positive residual (+2.24 log units), with actual revenue of €1.84B versus network-predicted €195M—a 9.4-fold gap. The GBA scenario reduced geriatrics’ residual to +1.52 (−0.72 change, 32% improvement) by transforming it from pure sink node to gateway node. The KHAG scenario produced negligible change (−0.002).

**Conclusions:** Network analysis reveals systematic structural-fiscal misalignment for geriatrics despite high service utilization. Strategic prerequisite placement achieves substantially greater alignment improvement than service group elimination. These findings demonstrate a quantitative approach for evaluating how regulatory frameworks recognize service importance, with potential implications for hospital planning in aging populations.

**Highlights:**

- Network analysis quantifies structural-fiscal alignment in German hospital reform
- 35.6% of service groups show good alignment between network position and revenue
- Geriatrics shows mathematical 9.4-fold gap: €1.84B actual vs. €195M predicted revenue
- Strategic prerequisite addition reduces alignment gap by 32% (in network-based predictions)
- Method provides generalizable framework for evaluating health policy reforms in terms of network metrics

## Introduction

Hospital reforms across developed nations increasingly structure accreditation and financing around defined service units or diagnosis groups with explicit quality standards and structural prerequisites [1–3]. The United Kingdom’s stroke service standards require co-located neurosurgery and intensive care in certified stroke units [4]. France’s perinatal networks mandate defined availability of obstetric and neonatal intensive care at referral centers [5]. These frameworks establish dependencies where certain services function as structural prerequisites enabling operation of others. Germany’s Krankenhausversorgungsverbesserungsgesetz (KHVVG; Hospital Care Improvement Act, enacted December 2024) represents a comprehensive extension of this approach, defining 65 hospital “Leistungsgruppen” (LGs, service groups) with codified mandatory and optional structural prerequisites across the entire hospital service system [6].

By design, these prerequisite relationships create a directed network structure. Some service groups function as enablers—required for operation of other services—while others remain structurally dependent. Since revenue is allocated at the LG level, this regulatory architecture may influence financial flows and hospital planning decisions. A critical but unanswered question is whether the network positions assigned by policy appropriately reflect the real-world economic importance of different services. Systematic misalignment could potentially create counterproductive incentives if network-based criteria increasingly influence allocation decisions, particularly for services crucial to demographic transitions but structurally peripheral in the regulatory framework.

Traditional health policy evaluation focuses on service volumes, costs, and clinical outcomes [3,7], but less attention has been given to the structural logic of reforms—that is, how legal definitions of prerequisites and dependencies shape systemic incentives. Network analysis provides a quantitative method to assess structural positioning [8,9]. In patient-sharing networks and referral systems, centrality metrics correlate with clinical influence and resource flows [10,11]. Applying similar methods to regulatory prerequisite networks could enable systematic comparison between policy-assigned structural positions and empirical measures of service importance.

Population aging fundamentally transforms healthcare needs [12,13]. Demand for services addressing multimorbidity, functional decline, and frailty grows rapidly as demographic pyramids invert [14,15]. Geriatrics emerged in the mid-20th century specifically to address complex needs of older adults [16,17], yet its institutional role within hospital systems remains contested [18]. In Germany, geriatrics represented €1.84 billion in hospital revenue in 2021, ranking among the top 10 service groups by economic volume [19]. However, preliminary analysis of the KHVVG dependency structure suggested geriatrics may occupy a peripheral network position despite this substantial utilization—raising concerns about structural-fiscal alignment as the population ages [6].

The KHVVG framework is not static. The current legislative draft (KHAG) proposes modifications including elimination of infectious diseases (LG3) and emergency medicine (LG65) as distinct service groups. Additionally, the Federal Joint Committee (Gemeinsamer Bundesausschuss, G-BA) has issued directives requiring geriatric co-management for specific surgical pathways, notably traumatic hip fractures requiring hip replacement [20]. These represent different intervention strategies: KHAG removes entire service nodes, while the G-BA directive adds prerequisite relationships between existing services or parts thereof. The comparative impact of these approaches on structural-fiscal alignment remains unexplored.

In this study, we applied network analytic methods to quantify structural-fiscal alignment in the KHVVG regulatory framework. We calculated centrality metrics for all 65 service groups and related these to empirical revenue data as a rough proxy for real-world service importance. We assessed the distribution of alignment across the system to validate the approach, with particular attention to geriatrics given its fiscal scale and relevance to demographic change. We then compared three policy scenarios (baseline KHVVG, the proposed KHAG modifications, and a hypothetical extension of G-BA geriatric prerequisites to all hip replacement services) to evaluate how different reform strategies affect structural-fiscal alignment.

Our aims were: (1) to demonstrate a quantitative method for assessing whether regulatory prerequisite structures reflect empirical service importance; (2) to characterize the distribution of structural-fiscal alignment across the German hospital system; (3) to identify services with substantial alignment gaps; and (4) to compare the effectiveness of different policy intervention strategies in addressing structural-fiscal misalignment.

## Materials and Methods

### Study Design

We conducted a quantitative network analysis of structural dependencies codified in the German KHVVG hospital reform. The reform defines 65 Leistungsgruppen (service groups) with formal prerequisites regarding presence or cooperation with other service groups. We modeled these prerequisites as a directed network to assess the relationship between policy-assigned structural position and empirical economic importance measured by service-specific revenue.

### Data Sources

Two primary data sources were used:

**Regulatory matrices:** Dependency relationships between LGs were extracted from official KHVVG legislative documentation. Each row encodes prerequisites for operating the respective service group; cells indicate whether another LG is required (mandatory vs. optional) and whether this requirement must be on-site or may be fulfilled through off-site cooperation.

**Revenue data:** Service group-level revenue data (full-year 2021, million EUR) were obtained from German hospital financial statistics and merged with LG identifiers as previously reported [19]. LG nomenclature was standardized from state-specific (Northrhine-Westphalia) to federal KHVVG numbering. Revenue serves as a proxy for real-world service utilization and economic importance, with explicitely acknowledged limitations that it reflects current reimbursement structures rather than clinical or societal value or population health impact.

### Policy Scenarios

Three scenarios were analyzed:

**Baseline (BASE):** Complete KHVVG structure with all 65 service groups and prerequisite relationships as codified in December 2024 legislation [6].

**KHAG scenario:** Modified structure reflecting proposed legislative changes that eliminate LG3 (Infektiologie, infectious diseases) and LG65 (Notfallmedizin, emergency medicine) as distinct service groups. This represents a node removal intervention.

**GBA scenario:** Hypothetical structure implementing extended geriatric prerequisites, modeled on G-BA directives for traumatic hip fracture but applied to all hip replacement services (LG23, Hip Endoprosthetics). Specifically, LG56 (Geriatrics) was added as a mandatory on-site prerequisite for LG23. This represents a strategic edge addition intervention. Due to data limitations, we could not distinguish traumatic from elective hip replacements as they are not reflected in the service group (Leistungsgruppen) structure.

### Network Modeling

Each LG was represented as a node in a directed graph. Prerequisites were modeled as directed edges from prerequisite LG to dependent LG. Edges were weighted to reflect legal strength:

- Mandatory on-site: 0.8
- Mandatory off-site: 0.4
- Optional on-site: 0.2
- Optional off-site: 0.1

To reduce combinatorial complexity, primary analyses used mandatory dependencies only (weights 0.8 and 0.4). Boolean logic encoded in prerequisite matrices (AND/OR groups) was implemented by preserving all AND-links and selecting one prerequisite per OR-clause through Monte Carlo sampling (10,000 iterations). The mean network realization across iterations was used for all analyses.

### Network Measures

For each LG, we calculated standard and adapted centrality metrics:

**Degree centrality:** In-degree (number of prerequisites required), out-degree (number of services depending on this LG), and total degree.

**Weighted degree:** Sum of edge weights for in-degree, out-degree, and total degree, reflecting both number and legal strength of connections.

**Enablement index:** Custom metric calculated as the weighted sum of outgoing mandatory edges adjusted for OR-clause fractions, quantifying the extent to which an LG functions as a structural prerequisite for other services.

**Betweenness centrality:** Fraction of shortest paths passing through the LG, indicating intermediary role in the dependency network.

**Closeness centrality:** Inverse of average distance to all other nodes, calculated separately for in-paths (as dependent) and out-paths (as enabler).

**PageRank:** Importance measure based on the link structure, adapted from web search algorithms.

**HITS scores:** Hub score (points to important nodes) and authority score (pointed to by important nodes).

**AND/OR clause counts:** Number of AND-type and OR-type prerequisite clauses referencing the LG, reflecting structural demand.

### Linking Network Position to Economic Importance

Revenue data were log-transformed (log1p) to address right-skewed distribution. Elastic net regression [21] related log-revenue to the full set of network metrics:

log(Revenue + 1) ~ α_0_ + Σj (αj × Network_Metric j) + 1

Elastic net combines L1 (lasso) and L2 (ridge) regularization to handle collinearity among network metrics while enabling feature selection. Regularization parameters (α, L1 ratio) were optimized via 5-fold cross-validation minimizing mean squared error. Models were fit separately for each policy scenario.

### Structural-Fiscal Alignment Assessment

The residual from regression quantifies alignment between network position and economic importance:

Residual = log(Actual Revenue + 1) − log(Predicted Revenue + 1)

**Positive residuals** indicate network metrics underpredict actual revenue, suggesting the service is more economically important than its structural position implies.

**Negative residuals** indicate network metrics overpredict revenue, suggesting the service is less important than its structural position implies.

**Near-zero residuals** (|residual| < 0.5 log units, corresponding to <65% prediction error) indicate good alignment between policy-assigned structure and empirical importance.

### Comparative Scenario Analysis

For each scenario (BASE, GBA, KHAG), we: 1. Reconstructed the dependency network, 2. Recalculated all network metrics for all 65 Lgs, 3. Re-fit elastic net regression models, 4. Computed residuals for all LGs and 5. Assessed changes in LG56 (Geriatrie) structural position and residual.

Model stability was assessed by correlating regression coefficients across scenarios using Pearson correlation.

### Software and Statistical Analysis

All analyses were conducted in Python 3.x with reproducible random seeds (seed=42). Network construction and graph-theoretical calculations used NetworkX (v2.x). Data handling utilized pandas and NumPy; regression modeling employed scikit-learn (ElasticNetCV with cross-validation). Correlation analyses used SciPy. Monte Carlo sampling for OR-clause resolution used custom implementations. Visualization employed Matplotlib. All code is available upon reasonable request.

### Ethical Considerations

All data were publicly available, aggregated at the service group level, and contained no individual patient information. No ethics approval was required.

## Results

### Baseline Network Structure

The KHVVG dependency network comprised 65 service groups (nodes) connected by 283 mandatory prerequisite relationships (directed edges). Mean weighted in-degree was 3.0 (SD 1.7), indicating most service groups depend on two to three mandatory prerequisites. Weighted out-degree distribution was highly right-skewed (median 0.40, IQR 0.00–1.40), with enablement index ranging from 0.0 to 45.2. Five service groups accounted for 48% of all mandatory outgoing edges: LG64 (Intensive Care, enablement index 45.2), LG1 (General Internal Medicine, 38.6), LG14 (General Surgery, 31.2), LG46 (Radiology, 5.2), and LG39 (Anesthesiology, 2.8).

### Network-Revenue Relationship

The elastic net regression model for the baseline scenario explained 56.6% of variance in log-transformed revenue (R²=0.566, optimized α=0.051, L1 ratio=0.50). Nine network metrics received non-zero coefficients. The strongest positive predictors were in-AND clauses (β=+0.61), enablement index (β=+0.38), HITS authority score (β=+0.14), and closeness centrality (in-direction, β=+0.11). The strongest negative predictors were weighted in-degree (β=−0.84) and PageRank (β=−0.40).

### Distribution of Structural-Fiscal Alignment

Across 60 service groups with available revenue data, residuals ranged from −3.00 to +2.24 log units. The distribution showed approximate normality around zero with extended tails. Twenty-one service groups (35.6%) had residuals within ±0.5 log units, indicating good alignment between network position and economic importance.

**Well-aligned major services** included: - LG1 (General Internal Medicine): Revenue €9,788M, residual +0.034 - LG14 (General Surgery): Revenue €12,128M, residual −0.059 - LG20 (Complex Peripheral Arterial Vessels): Revenue €626M, residual −0.022 - LG36 (Ophthalmology): Revenue €481M, residual +0.071 - LG60 (Lung Transplantation): Revenue €46M, residual +0.074

**Services with large positive residuals** (network position undervalues economic importance): - LG56 (Geriatrics): Revenue €1,843M, predicted €195M, residual +2.24 - LG28 (Spinal Surgery): Revenue €1,622M, predicted €282M, residual +1.75 - LG7 (Pediatric Surgery): Revenue €164M, predicted €32M, residual +1.62 - LG54 (Stroke Unit): Revenue €1,103M, predicted €228M, residual +1.57 - LG42 (Orthopedics/Trauma): Revenue €1,766M, predicted €376M, residual +1.54

**Services with large negative residuals** (network position overvalues economic importance): - LG58 (Intestinal Transplantation): Revenue €1M, predicted €39M, residual −3.00 - LG63 (Pancreatic Transplantation): Revenue €2M, predicted €39M, residual −2.59 - LG31 (Heart Transplantation): Revenue €111M, predicted €590M, residual −1.66 - LG30 (Lung Transplantation): Revenue €137M, predicted €638M, residual −1.53 - LG40 (Complex Cardiac Surgery): Revenue €79M, predicted €356M, residual −1.50

### Baseline Position of LG56 (Geriatrics)

In the baseline KHVVG matrix, LG56 (Geriatrics) exhibited the largest positive residual among all 60 service groups with revenue data. Network metrics: weighted in-degree 4.8, weighted out-degree 0.0, enablement index 0.0, betweenness centrality 0.0, PageRank 0.033. The regression model predicted log-revenue of 5.28, corresponding to €195M. Actual revenue was €1,843M (log-revenue 7.52), yielding a residual of +2.24 log units. The actual-to-predicted ratio was 9.4-fold.

### Comparative Scenario Analysis: Network Metrics

**GBA scenario** (adding LG56 as prerequisite for LG23 / Hip Endoprosthetics):

LG56 network metrics changed as follows: - Degree: in-degree 10→10, out-degree 0→1, total degree 10→11 - Weighted degree: in 4.8→4.8, out 0.0→0.4, total 4.8→5.2 - Enablement index: 0.0→0.4 - Betweenness centrality: 0.0→0.012 - Closeness (in): 0.0→0.028 - PageRank: 0.033→0.039 (+20%)

**KHAG scenario** (removing LG3 and LG65):

LG56 network metrics changed as follows: - Degree: in-degree 10→10, out-degree 0→0, total degree 10→10 - Weighted degree: in 4.8→4.8, out 0.0→0.0, total 4.8→4.8 - Enablement index: 0.0→0.0 - Betweenness centrality: 0.0→0.0 - Closeness (in): 0.0→0.0 - PageRank: 0.033→0.034 (+2%)

### Comparative Scenario Analysis: Revenue Predictions and Residuals

**BASE scenario:** - LG56 predicted revenue: €195M - LG56 actual revenue: €1,843M - Residual: +2.244 log units

**GBA scenario:** - LG56 predicted revenue: €401M - LG56 actual revenue: €1,843M (unchanged, fixed empirical benchmark) - Residual: +1.522 log units - Change from BASE: −0.722 log units (32% reduction in alignment gap)

**KHAG scenario:** - LG56 predicted revenue: €195M - LG56 actual revenue: €1,843M (unchanged, fixed empirical benchmark) - Residual: +2.242 log units - Change from BASE: −0.002 log units (0.1% change)

### Model Stability Across Scenarios

Elastic net models for all three scenarios showed similar performance: - BASE: R²=0.566, 9 non-zero coefficients - GBA: R²=0.571, 11 non-zero coefficients

- KHAG: R²=0.566, 9 non-zero coefficients

Regression coefficients showed high correlation between scenarios: - BASE vs. KHAG: Pearson r=1.00 - BASE vs. GBA: Pearson r=0.94

Major coefficient changes from BASE to GBA: - Weighted in-degree: −0.84 to −0.55 (34% reduction in magnitude) - In-AND clauses: +0.61 to +0.43 (−29%) - Enablement index: +0.38 to +0.26 (−33%) - Betweenness: +0.072 to +0.39 (+447%)

All coefficients in KHAG remained within 1% of BASE values for the top predictors (weighted in-degree, in-AND clauses, enablement index, PageRank).

### Distribution of Alignment Changes Across All Service Groups

In the GBA scenario, 58 of 60 service groups (97%) experienced residual changes <0.1 log units. Only LG56 (Geriatrics, −0.72) and LG23 (Hip Endoprothetics, +0.44) showed substantial changes. In the KHAG scenario, all 60 service groups experienced residual changes <0.01 log units, with mean absolute change of 0.001.

## Discussion

### Principal Findings

This study demonstrates the first quantitative approach for assessing the structural-fiscal alignment in regulatory frameworks for hospital services in Germany. Applying network analysis to Germany’s KHVVG reform, we found that 35.6% of service groups showed good alignment between network-assigned structural position and empirical economic importance. However, geriatrics exhibited the largest misalignment: network metrics predicted only €195M revenue versus actual €1,843M - a 9.4-fold gap. Comparative scenario analysis revealed markedly asymmetric policy effectiveness: strategic prerequisite addition (GBA scenario) reduced the geriatrics alignment gap by 32%, while service group elimination (KHAG scenario) produced <1% change. These findings have potential implications for hospital planning in aging populations and for the design of prerequisite-based regulatory frameworks more broadly.

The finding that over one-third of service groups showed good structural-fiscal alignment validates the analytical approach. Major services with well-aligned network positions included general internal medicine (€9.8B, residual +0.03), general surgery (€12.1B, residual −0.06), and complex peripheral vascular services (€626M, residual −0.02). These services function as central hubs in the prerequisite network and generate revenue consistent with their structural importance. The regression model’s explanatory power (R²=0.566) indicates that network position meaningfully predicts economic importance for most services, while substantial variance remains, which is to be expected, as revenue reflects factors beyond structural dependencies including disease prevalence, treatment costs, and reimbursement rates.

Services with large negative residuals (transplantation specialties) generate less revenue than their central network positions would predict, likely reflecting low case volumes despite high structural requirements. Services with large positive residuals generate more revenue than structural position suggests, indicating either high-volume utilization not captured by prerequisite relationships or structural undervaluation in the regulatory framework. The existence of both well-aligned and misaligned services demonstrates that the method discriminates meaningfully rather than finding systematic over- or underprediction.

### Structural-Fiscal Misalignment in Geriatrics

Geriatrics’ position as the most structurally peripheral service group is notable on multiple grounds. First, the magnitude: a 9.4-fold gap represents the largest alignment discrepancy in the system.

Second, the absolute scale: €1.84 billion in annual revenue places geriatrics among the top 10 service groups economically, yet its network position (weighted in-degree 4.8, enablement index 0.0) is peripheral. Third, the demographic context: with Germany’s population aged ≥65 projected to reach 28% by 2035, services addressing multimorbidity and frailty face increasing demand [22,23].

The structural-fiscal discordance arises from geriatrics’ classification in the KHVVG framework: it requires multiple prerequisites (general internal medicine, diagnostics, etc.) but serves as a mandatory prerequisite for no other services. This creates a pure sink node in the dependency network. The regression model identifies enablement index as a strong positive predictor (β=+0.38) and weighted in-degree as a strong negative predictor (β=−0.84), penalizing services that depend on others while not enabling others. Geriatrics’ profile thus would predict low revenue, contradicting empirical reality.

This misalignment is a mathematical finding primarily, but it will matter if resource allocation mechanisms increasingly weight network-based criteria, e.g. if flow-based approaches are proposed [22]. Current financing relies on diagnosis-related groups [1], but policy discussions also can emphasize value-based care, bundled payments, and quality incentives [23] - all potentially influenced by structural positioning. If prerequisite status signals strategic importance in allocation decisions, services with high dependency but low enablement may face structural challenges despite high utilization.

### Comparative Effectiveness of Policy Interventions

The GBA scenario added a single prerequisite edge: geriatrics became mandatory for hip replacement (LG23). This minimal structural change substantially improved alignment, reducing the residual from +2.24 to +1.52 (−0.72, 32% improvement), while changing virtually nothing for the other LGs. The mechanism: LG56’s enablement index increased from 0.0 to 0.4, betweenness centrality from 0.0 to 0.012, and PageRank by 20%. These centrality increases shifted predicted revenue from €195M to €401M, thus more than doubling the network-based valuation.

The clinical plausibility of this intervention strengthens its policy relevance. Hip fracture patients frequently present with multimorbidity, functional decline, and delirium [24]. Geriatric co-management for hip fracture reduces complications, length of stay, and mortality in multiple randomized trials [25–27]. The G-BA has mandated geriatric consultation for traumatic hip fractures requiring replacement [19]. Our GBA scenario extends this logic to all hip replacement, acknowledging that elective hip replacement might not encounter similar patient populations. Still, data limitations prevent us from performing more fine-grained adjustments.

In contrast, the KHAG scenario removed two entire service groups (infectious diseases, emergency medicine) but produced negligible impact on geriatrics’ alignment (−0.002 residual change, <1%). Neither removed service was a direct prerequisite for geriatrics, nor did geriatrics serve as prerequisite for them. The removal occurred outside geriatrics’ local network neighborhood, leaving its centrality metrics essentially unchanged. All 60 service groups experienced <0.01 residual change in KHAG, indicating the removed services were relatively isolated in the dependency network in the first place.

These contrasting results demonstrate that policy effectiveness depends critically on where interventions occur in the network structure. Strategic edge placement in high-traffic pathways (hip endoprosthetics → geriatrics) generates disproportionate effects, while node removal in peripheral regions produces minimal impact.

### Potential Implications for Hospital Planning

From a hospital planning perspective, structural-fiscal misalignment might create asymmetric incentives. Services with strong enabling roles (high enablement index) are structurally central or up-stream: many other services depend on them, potentially signaling strategic importance in allocation decisions. Services with high dependency but low enablement face structurally peripheral placement despite potentially high demand. For geriatrics specifically, structural-fiscal misalignment could discourage capacity expansion or investment despite demographic need, if decision-makers rely on structural signals from regulatory frameworks.

The GBA scenario demonstrates that targeted prerequisite additions can improve alignment. However, indiscriminate addition of prerequisites risks overbuilding network density, potentially pushing the system toward hospital “super-clusters” where every facility must house all services—an inefficient and undesirable outcome. The appropriate balance likely involves selective prerequisite relationships that reflect documented clinical pathways and care dependencies.

An important caveat: our analysis models structural prerequisites, not actual patient flows. While network closeness may facilitate clinical workflows, these concepts are not equivalent. Upgrading every high-revenue service’s structural position to match economic importance would artificially inflate centrality without necessarily improving care coordination. The goal should be alignment where mismatches create demonstrable problems, and where mismatches are highly unbalanced, but not mechanical matching of structure to revenue.

### Broader Implications for Health System Design

Beyond the German context, these findings illustrate a generalizable approach for evaluating prerequisite-based regulatory frameworks. Network analysis enables systematic assessment of whether dependencies contained therein align with empirical utilization patterns. Large misalignments might serve as an indicator warranting policy attention: either the structural requirements fail to capture actual care pathways (suggesting prerequisite modification), or revenue patterns reflect inadequate incentives unrelated to clinical need (suggesting reimbursement reform).

The method also reveals intervention leverage points. Not all policy changes equally affect structural-fiscal alignment. Strategic edge modifications in high-centrality regions will naturally achieve greater effect than bulk node elimination, particularly when removed nodes occupy peripheral positions.

### Limitations

Several limitations merit consideration. First, revenue is an imperfect proxy for service importance. It reflects current reimbursement structures, case volumes, and cost structures and not necessarily clinical value, quality, or population health impact. Services with high revenue but poor outcomes would appear “important” by this metric. Also, we cannot assess whether the current revenue patterns represent optimal or appropriate service utilization. Nevertheless, revenue numbers do signal actual real-world utilization, and systematic misalignment between structural position and utilization creates planning challenges regardless of normative judgments about appropriate utilization levels.

Second, our analysis is cross-sectional, precluding causal inference. We cannot determine whether KHVVG implementation will change revenue patterns, only whether the structure aligns with current patterns. Longitudinal analysis post-implementation would enable causal assessment of structural effects on resource flows.

Third, the GBA scenario is hypothetical. While based on actual G-BA directives for traumatic hip fracture, we modeled application to all hip replacements due to data limitations preventing distinction of traumatic versus elective cases. The actual policy impact will therefore differ from our projection.

Fourth, we restricted analysis to mandatory prerequisites (weights 0.8, 0.4) to manage computational complexity. Optional relationships and additional possibilities contained in legislative exceptions may influence actual clinical networks. However, several robustness analyses across weight schemes and thresholds showed consistent patterns [data not shown], suggesting findings are not artifacts of specific threshold choices.

Fifth, model stability was very high (r=1.00 for BASE vs. KHAG), which while demonstrating robustness, also indicates the KHAG intervention was too small to substantively test model assumptions. The GBA scenario showed more coefficient variation (r=0.94), suggesting greater structural reorganization.

Finally, our focus on a single service group (Geriatrics) as primary case study limits generalizability. Other services with large residuals (Spinal Surgery +1.75, Pediatric Surgery +1.62, Stroke Units +1.57) warrant similar detailed analysis but were beyond this study’s scope.

## Conclusion

Network analysis provides a transparent method for quantifying structural-fiscal alignment in regulatory frameworks for hospital services. In Germany’s KHVVG reform, most major services show appropriate alignment, but geriatrics exhibits a 9.4-fold gap between network-predicted and actual economic importance. Strategic prerequisite placement substantially reduces this misalignment, while service group elimination produces negligible effect.

These findings could have immediate relevance for German hospital policy as KHAG modifications are debated. More broadly, the approach demonstrates how graph-theoretical methods can evaluate whether health system regulations appropriately recognize service importance, particularly critical for specialties serving aging populations where structural undervaluation could have long-term consequences for access, capacity and finally the quality of our health care system.

**Figure 1.**
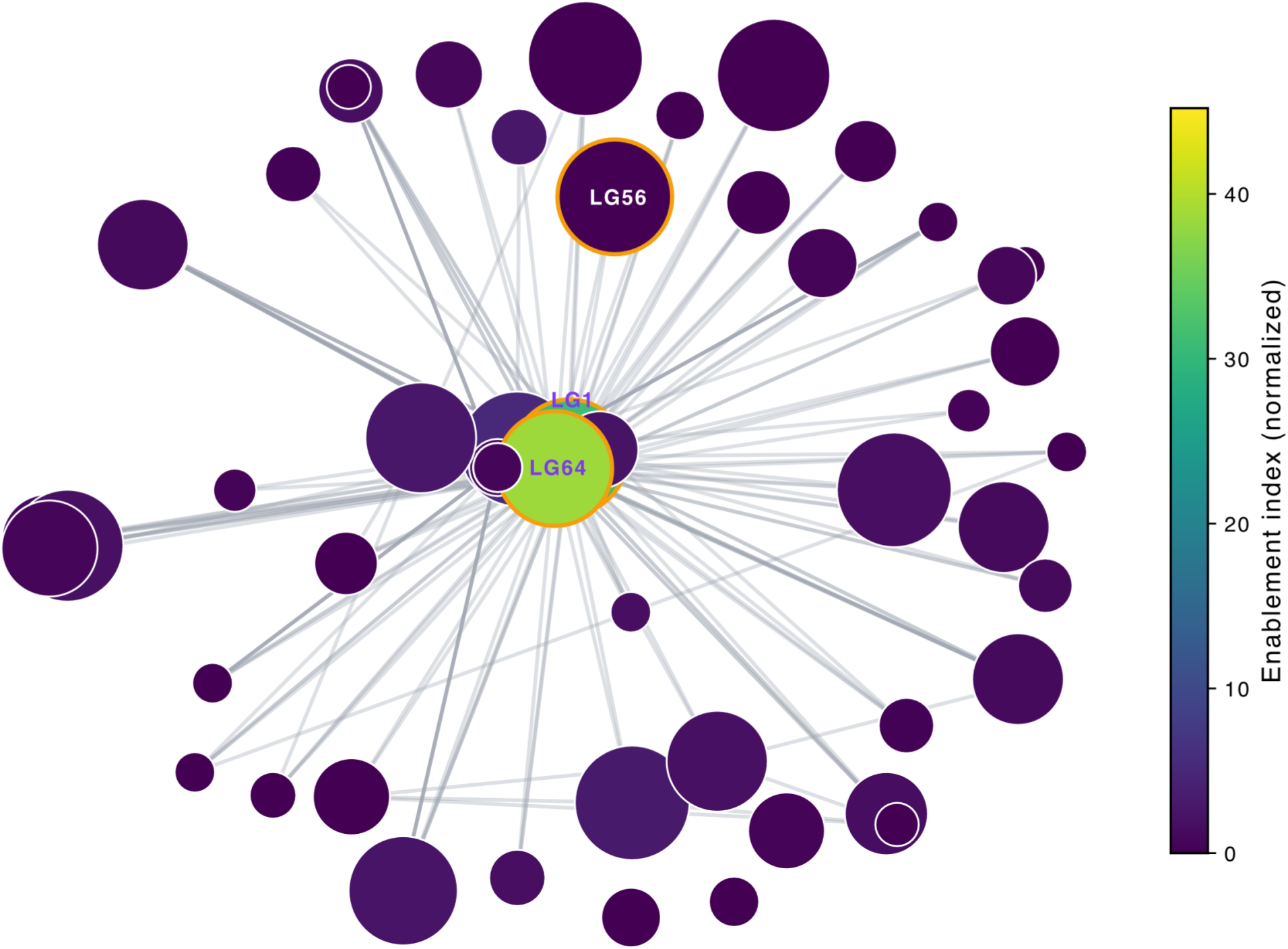
KHVVG Dependency Network Structure (BASE Scenario) Directed network graph showing mandatory prerequisite relationships (edge weight ≥0.4) among 65 hospital service groups. Nodes represent individual service groups; directed edges indicate prerequisite dependencies (arrow points from prerequisite to dependent service). Node size is proportional to total revenue (log scale). Node color represents enablement index (scale: blue=0 to yellow=45.2). Selected service groups are labeled: LG1 (General Internal Medicine), LG56 (Geriatrics), LG64 (Intensive Care). Layout uses force-directed algorithm (Fruchterman-Reingold) to position highly connected nodes centrally.

**Figure 2.**
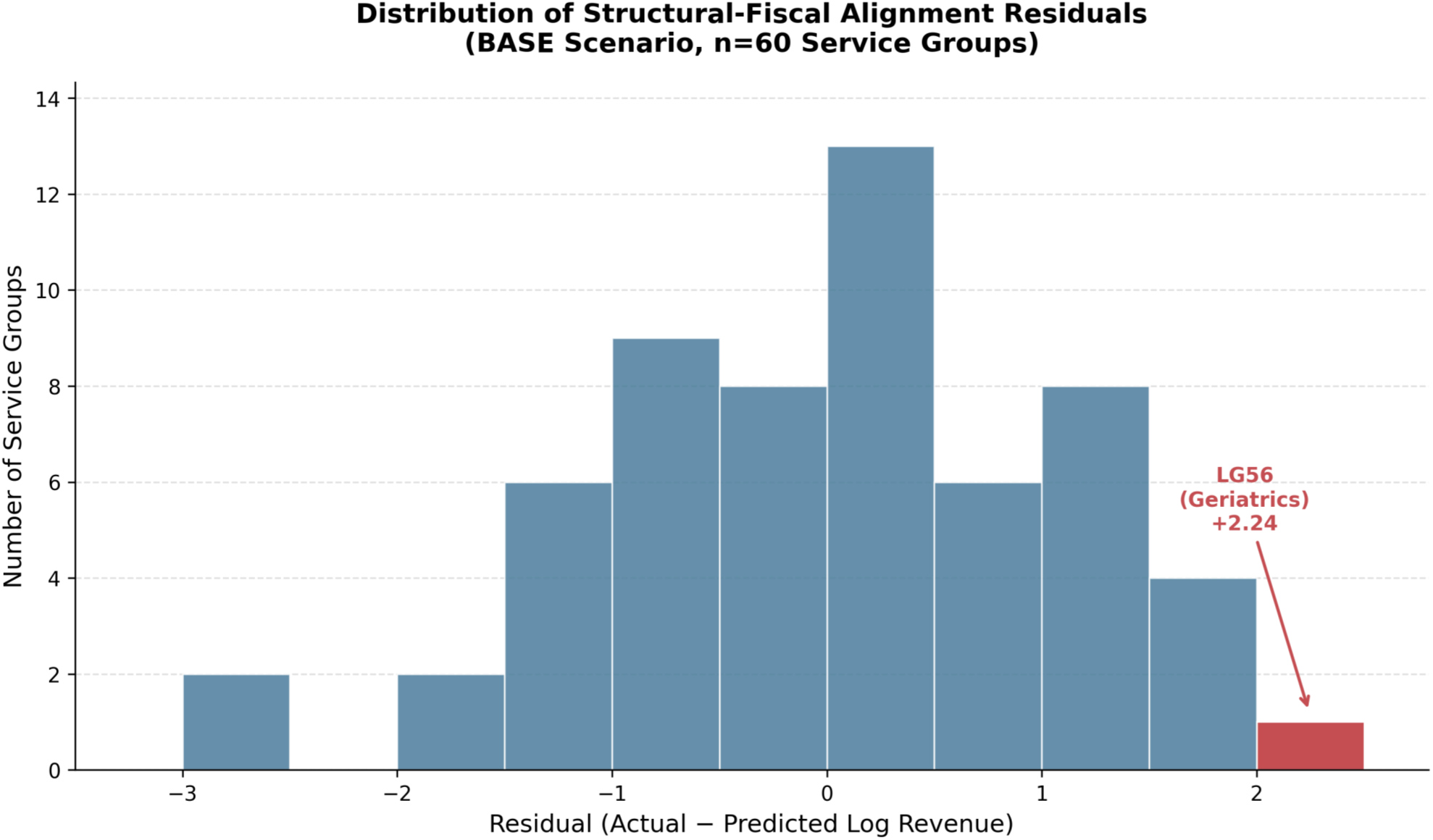
Distribution of Structural-Fiscal Alignment Across Service Groups. Histogram showing distribution of prediction residuals (actual minus predicted log-revenue) for all 60 service groups with revenue data (BASE scenario). Residuals are grouped in 0.5 log-unit bins. The distribution shows approximate normality. LG56 (Geriatrics) is highlighted in red.

**Figure 3.**
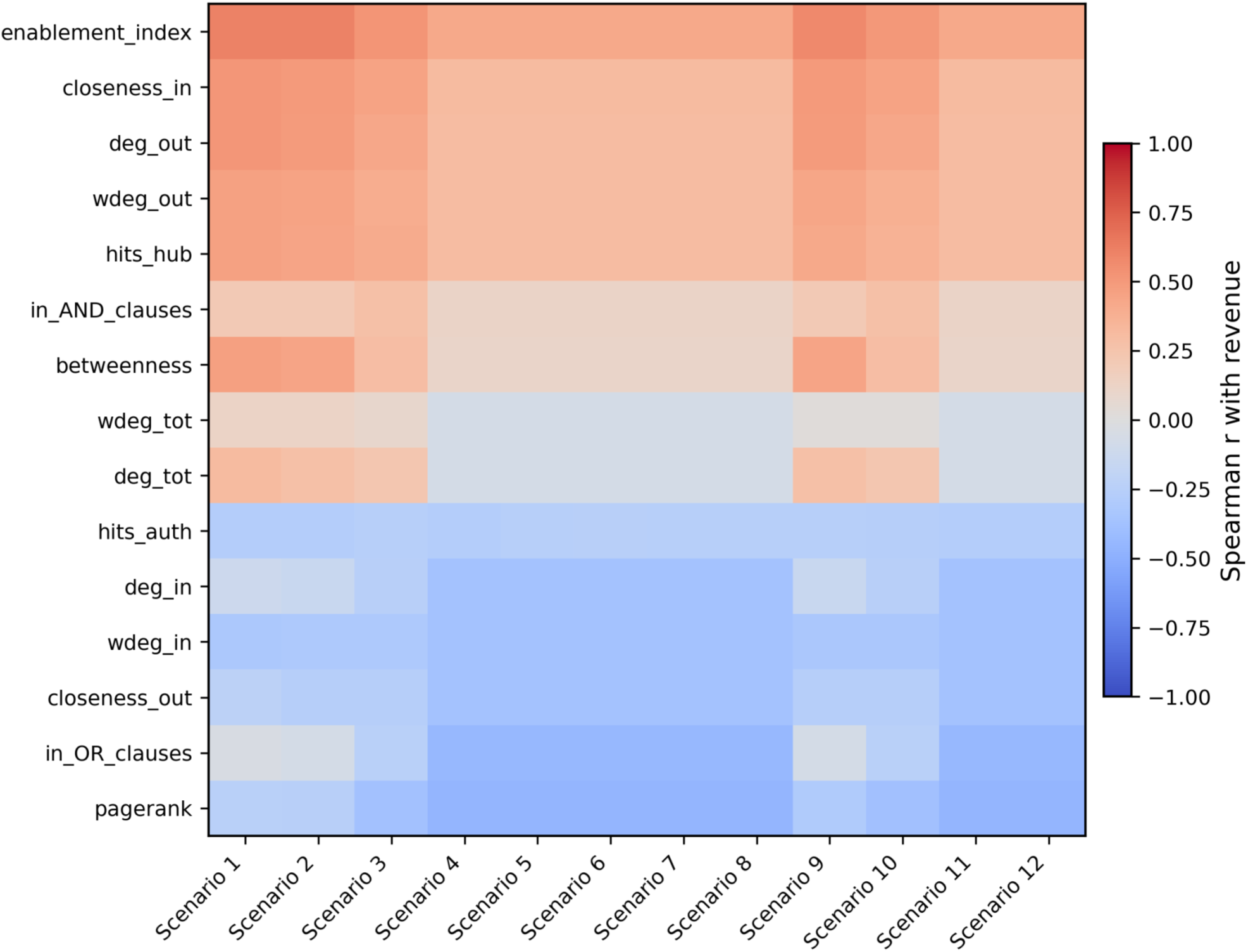
Model Stability and Coefficient Changes Across Network Configurations. Robustness heatmap of Spearman correlations between structural metrics and revenue across alternative network definitions using different OR connections, respectively, within the BASE scenario. Each column represents a scenario (Scenario 1–12), and each row a network metric. Blue shading indicates negative correlations, red shading positive correlations. Consistently positive correlations for enablement-related measures and negative correlations for dependency load demonstrate the stability of elastic net’s findings across specifications.

**Table 1.**
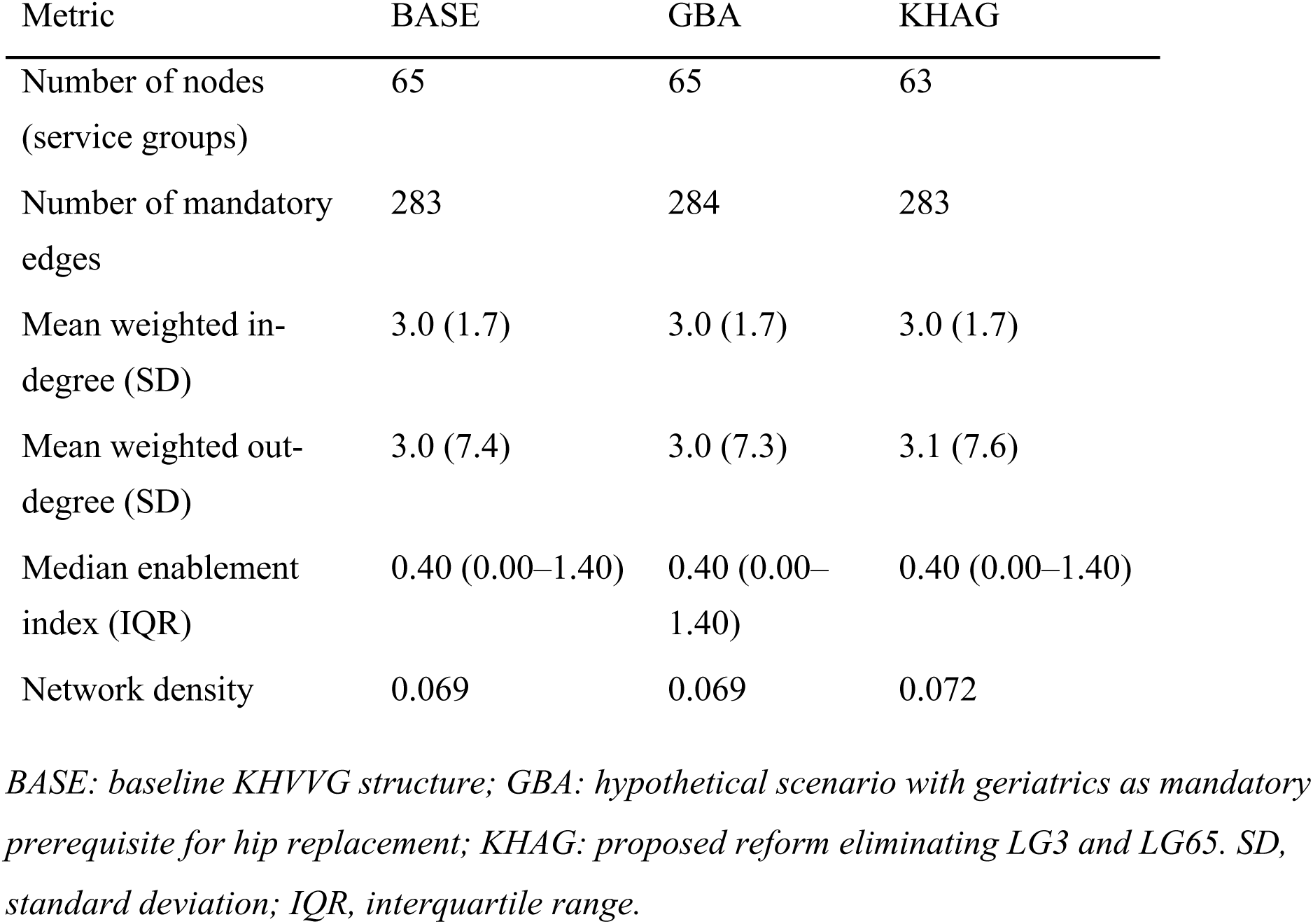
Network Structure Characteristics Across Policy Scenarios.

| Metric | BASE | GBA | KHAG |
| --- | --- | --- | --- |
| Number of nodes<br>(service groups) | 65 | 65 | 63 |
| Number of mandatory<br>edges | 283 | 284 | 283 |
| Mean weighted in-degree (SD) | 3.0 (1.7) | 3.0 (1.7) | 3.0 (1.7) |
| Mean weighted out-degree (SD) | 3.0 (7.4) | 3.0 (7.3) | 3.1 (7.6) |
| Median enablement<br>index (IQR) | 0.40 (0.00–1.40) | 0.40 (0.00–<br>1.40) | 0.40 (0.00–1.40) |
| Network density | 0.069 | 0.069 | 0.072 |

*BASE: baseline KHVVG structure; GBA: hypothetical scenario with geriatrics as mandatory prerequisite for hip replacement; KHAG: proposed reform eliminating LG3 and LG65. SD, standard deviation; IQR, interquartile range.*

**Table 2.** Elastic Net Regression Model Performance and Primary Predictors.

| Scenario | R<sup>2</sup> | Non-zero Coefficients | Top Positive Predictors ($\beta$) | Top Negative Predictors ($\beta$) |
| --- | --- | --- | --- | --- |
| **BASE** | 0.566 | 9 | in-AND clauses (+0.61)<br>Enablement index (+0.38)<br>HITS authority (+0.14)<br>Closeness (in) (+0.11) | Weighted in-degree (−0.84)<br>PageRank (−0.40) |
| **GBA** | 0.571 | 11 | Betweenness (+0.39)in-AND clauses (+0.43)<br>Enablement index (+0.26)<br>Closeness (in) (+0.20) | Weighted in-degree (−0.55)<br>PageRank (−0.29) |
| **KHAG** | 0.566 | 9 | in-AND clauses (+0.61)<br>Enablement index (+0.38)<br>HITS authority (+0.13)<br>Closeness (in) (+0.11) | Weighted in-degree (−0.83)<br>PageRank (−0.40) |

*All models used 5-fold cross-validation to optimize regularization parameters. Coefficient correlation: BASE vs. KHAG $r=1.00$ ; BASE vs. GBA $r=0.94$ .*

**Table 3.**
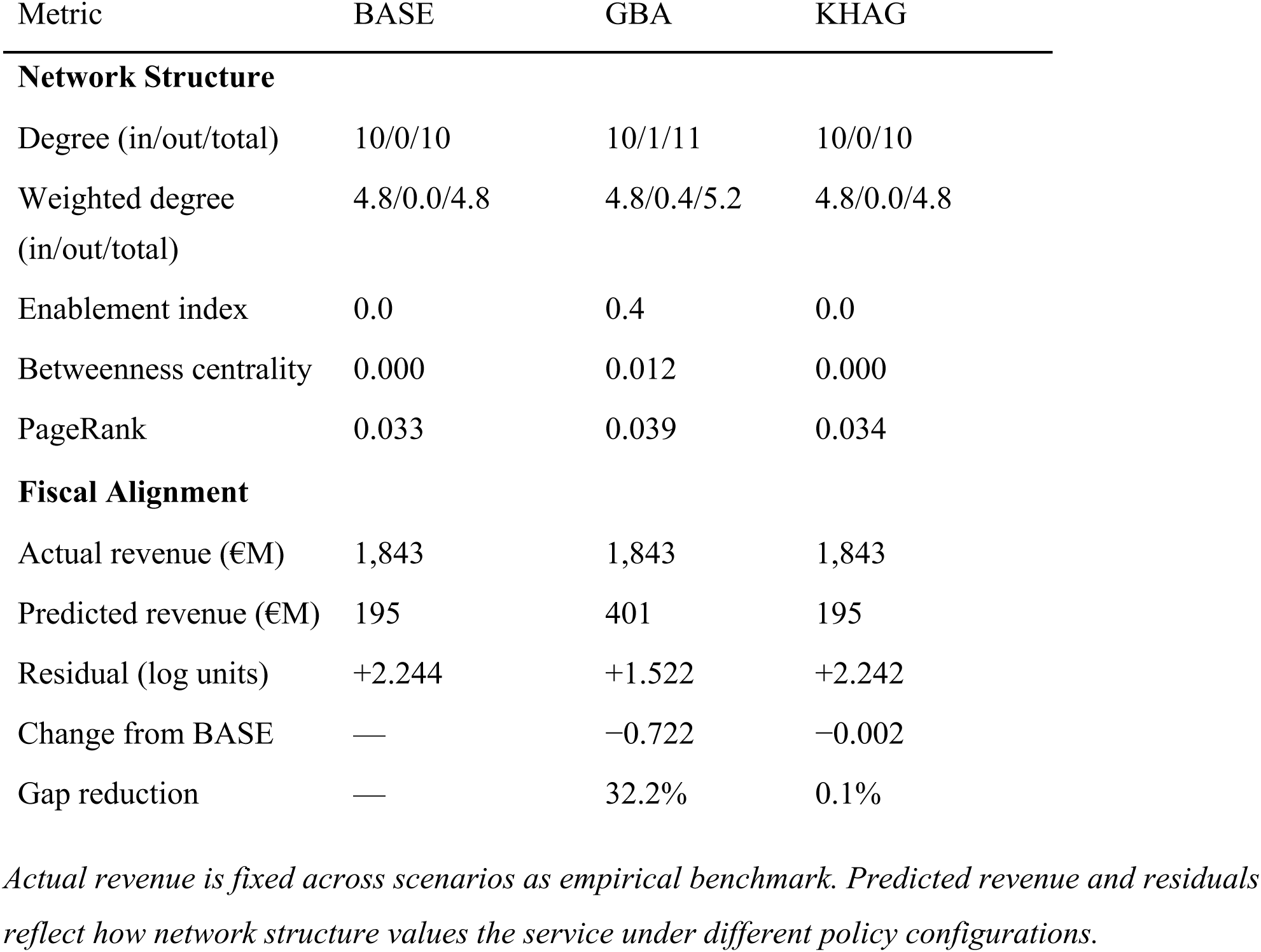
Network Metrics and Fiscal Alignment for LG56 (Geriatrics) Across Policy Scenarios.

**Table 4.** Representative Service Groups by Structural-Fiscal Alignment Category (BASE Scenario)

| Alignment Category | LG Code | Service Group | Revenue (€M) | Predicted (€M) | Residual | Weighted Degree (in/out) |
| --- | --- | --- | --- | --- | --- | --- |
| **Structurally**<br>**Concordant** | LG1 | General Internal Medicine | 9,788 | 9,458 | +0.03 | 40.8/38.6 |
|  | LG14 | General Surgery | 12,128 | 12,860 | −0.06 | 32.4/31.2 |
|  | LG20 | Complex Peripheral Vessels | 626 | 640 | −0.02 | 6.8/2.3 |
| **Structurally**<br>**peripheral,**<br>**revenue-rich**<br>(positive residuals) | LG56 | Geriatrics | 1,843 | 195 | +2.24 | 4.8/0.0 |
|  | LG28 | Spinal Surgery | 1,622 | 282 | +1.75 | 2.4/0.0 |
|  | LG54 | Stroke Unit | 1,103 | 228 | +1.57 | 5.2/0.7 |
| **Structurally**<br>**central,**<br>**revenue-poor**<br>(negative residuals) | LG58 | Intestinal Transplantation | 1 | 39 | −3.00 | 6.4/0.0 |
|  | LG63 | Pancreatic Transplantation | 2 | 39 | −2.59 | 6.4/0.0 |
|  | LG31 | Heart Transplantation | 111 | 590 | −1.66 | 6.8/0.6 |

*Structural-fiscal concordance: |residual| < 0.5 log units. Structural-fiscal discordance: |residual| ≥ 0.5 log units; network position under- (positive residual) or overvalues (negative residual) economic importance.*

## Data Availability

All data produced in the present study are available upon reasonable request to the authors.

## References

[1] Busse R, Geissler A, Aaviksoo A, et al. Diagnosis related groups in Europe: moving towards transparency, efficiency, and quality in hospitals? BMJ 2013;346:f3197. 10.1136/bmj.f3197.

[2] Geissler A, Scheller-Kreinsen D, Quentin W, et al. Do Diagnosis-Related Groups Appropriately Explain Variations in Costs and Length of Stay of Hip Replacement? A Comparative Assessment of Drg Systems Across 10 European Countries. Health Econ 2012;21:103–15. 10.1002/hec.2848.

[3] Kimberly J, De Pouvourville G, d’Aunno T, editors. The Globalization of Managerial Innovation in Health Care. 1st ed. Cambridge University Press; 2001. 10.1017/CBO9780511620003.

[4] NHS England. National Stroke Service Model: Integrated Stroke Delivery Networks. NHS England and NHS Improvement; 2021.

[5] Ville Y, Rudigoz RC, Hascoët JM. Rapport 23-05. Planification d’une politique en matière de périnatalité en France: organiser la continuité des soins est une nécessité et une urgence. Bull Académie Natl Médecine 2023;207:560–75. 10.1016/j.banm.2023.03.017.

[6] Bundesministerium für Gesundheit. Krankenhausversorgungsverbesserungsgesetz (KHVVG). 2024.

[7] Palmer K, Marengoni A, Forjaz MJ, et al. Multimorbidity care model: Recommendations from the consensus meeting of the Joint Action on Chronic Diseases and Promoting Healthy Ageing across the Life Cycle (JA-CHRODIS). Health Policy 2018;122:4–11. 10.1016/j.healthpol.2017.09.006.

[8] Korsberg A, Cornelius SL, Awa F, et al. A Scoping Review of Multilevel Patient-Sharing Network Measures in Health Services Research. Med Care Res Rev MCRR 2025;82:203–24. 10.1177/10775587241304140.

[9] Beaney T, Clarke J, Barahona M, et al. A primary care network analysis: natural communities of general practices in London. Br J Gen Pract J R Coll Gen Pract 2020;70:bjgp20X711113. 10.3399/bjgp20X711113.

[10] Breslau J, Dana B, Pincus H, et al. Empirically identified networks of healthcare providers for adults with mental illness. BMC Health Serv Res 2021;21:777. 10.1186/s12913-021-06798-2.

[11] Landon BE, Keating NL, Barnett ML, et al. Variation in Patient-Sharing Networks of Physicians Across the United States. JAMA 2012;308:265–73. 10.1001/jama.2012.7615.

[12] Skou ST, Mair FS, Fortin M, et al. Multimorbidity. Nat Rev Dis Primer 2022;8:48. 10.1038/s41572-022-00376-4.

[13] Vetrano DL, Palmer K, Marengoni A, et al. Frailty and Multimorbidity: A Systematic Review and Meta-analysis. J Gerontol Ser A 2019;74:659–66. 10.1093/gerona/gly110.

[14] Rodrigues LP, de Oliveira Rezende AT, Delpino FM, et al. Association between multimorbidity and hospitalization in older adults: systematic review and meta-analysis. Age Ageing 2022;51:afac155. 10.1093/ageing/afac155.

[15] Rechel B, Grundy E, Robine J-M, et al. Ageing in the European Union. The Lancet 2013;381:1312–22. 10.1016/S0140-6736(12)62087-X.

[16] Ritch A. History of geriatric medicine: from Hippocrates to Marjory Warren. J R Coll Physicians Edinb 2012;42:368–74. 10.4997/JRCPE.2012.417.

[17] Evans JG. Geriatrics. Postgrad Med J 2003;79:245–50.

[18] Bardach SH, Rowles GD. Geriatric Education in the Health Professions: Are We Making Progress? The Gerontologist 2012;52:607–18. 10.1093/geront/gns006.

[19] Schmitt J, Sundmacher L, Augurzky B, et al. Krankenhausreform in Deutschland: Populationsbezogenenes Berechnungs- und Simulationsmodell zur Planung und Folgenabschätzung. Monit Versorg 2024;2024:37–50. 10.24945/MVF.03.24.1866-0533.2606.

[20] Bundesministerium für Gesundheit. Beschluss des Gemeinsamen Bundesausschusses über eine Richtlinie zur Versorgung der hüftgelenknahen Femurfraktur. 2020.

[21] Zou H, Hastie T. Regularization and Variable Selection Via the Elastic Net. J R Stat Soc Ser B Stat Methodol 2005;67:301–20. 10.1111/j.1467-9868.2005.00503.x.

[22] Rechel B, Wright S, Barlow J, et al. Hospital capacity planning: from measuring stocks to modelling flows. Bull World Health Organ 2010;88:632–6. 10.2471/BLT.09.073361.

[23] Porter ME, Lee TH. The Strategy That Will Fix Health Care. Harv Bus Rev n.d.

[24] Penrod JD, Boockvar KS, Litke A, et al. Physical Therapy and Mobility 2 and 6 Months After Hip Fracture. J Am Geriatr Soc 2004;52:1114–20. 10.1111/j.1532-5415.2004.52309.x.

[25] Folbert EC, Hegeman JH, Vermeer M, et al. Improved 1-year mortality in elderly patients with a hip fracture following integrated orthogeriatric treatment. Osteoporos Int 2017;28:269–77. 10.1007/s00198-016-3711-7.

[26] Grigoryan KV, Javedan H, Rudolph JL. Orthogeriatric care models and outcomes in hip fracture patients: a systematic review and meta-analysis. J Orthop Trauma 2014;28:e49–55. 10.1097/bot.0b013e3182a5a045.

[27] Prestmo A, Hagen G, Sletvold O, et al. Comprehensive geriatric care for patients with hip fractures: a prospective, randomised, controlled trial. The Lancet 2015;385:1623–33. 10.1016/S0140-6736(14)62409-0.

